# Predicting Suicidality in people living with HIV in Uganda: A Machine Learning Approach

**DOI:** 10.1101/2025.03.06.25323536

**Authors:** Anthony B. Mutema, Linda Lillian, Daudi Jjingo, Segun Fatumo, Eugene Kinyanda, Allan Kalungi

**Affiliations:** African Center of Excellence in Bioinformatics and Data Intensive Science, Makerere University, Kampala, Uganda; AIDS Healthcare Foundation (AHF-Uganda Cares), Kampala, Uganda; Medical Research Council/Uganda Virus Research Institute and London School of Hygiene and Tropical Medicine (MRC/UVRI and LSHTM) Uganda Research Unit, Entebbe, Uganda; Precision Healthcare University Research Institute, Queen Mary University of London, London, UK; Department of Non-communicable Disease Epidemiology, London School of Hygiene and Tropical Medicine, London, UK

**Keywords:** Suicidality, prediction, machine learning, polygenic risk scores, HIV

## Abstract

**Background:** People living with HIV (PLWH) are more likely to experience suicidal thoughts and exhibit suicidal behavior than the general population. However, there are currently no effective methods of predicting who is likely to experience suicidal thoughts and behavior. Machine learning (ML) approaches can be leveraged to develop models that evaluate the complex etiology of suicidal behavior, facilitating the timely identification of at-risk individuals and promoting individualized treatment allocation.

**Materials and methods:** This retrospective case-control study used longitudinal sociodemographic, psychosocial, and clinical data of 1,126 PLWH from Uganda to evaluate the potential of ML in predicting suicidality. In addition, suicidality polygenic risk scores (PRS) were calculated for a subset of 282 study participants and incorporated as an additional feature in the model to determine if including genomic information improves overall model performance. The model’s performance was evaluated using the area under the receiver operating characteristics curve (AUC), positive predictive value (PPV), sensitivity, specificity, and Mathew’s correlation coefficient (MCC).

**Results:** We trained and evaluated eight different ML algorithms including logistic regression, support vector machines, Naïve Bayes, k-nearest neighbors, decision trees, random forests, AdaBoost, and gradient-boosting classifiers. Cost-sensitive AdaBoost emerged as the best model, achieving an AUC of 0.79 (95% CI: 0.72–0.87), a sensitivity of 0.63, a specificity of 0.74, a PPV of 0.36, and an NPV of 0.89 on unseen baseline data. The model demonstrated good generalizability, predicting prevalent and incident suicidality at 12-month follow-up with an AUC of 0.75 (95% CI: 0.69–0.81) and 0.69 (95% CI: 0.62–0.76) respectively. Incorporating PRS as an additional feature in the model resulted in a 19% and 14% improvement in model sensitivity and PPV respectively, and a 4% reduction in specificity. A positive MDD diagnosis and high stress contributed the most to predicting suicidality risk.

**Conclusion:** A cost-sensitive AdaBoost model developed using the sociodemographic, psychosocial, and clinical data of PLWH in Uganda can predict suicidality risk. Incorporating suicidality PRS improved the overall predictive performance of the model. However, larger studies involving more diverse participants are needed to evaluate the potential of PRS in enhancing risk stratification and the clinical utility of the prediction model.

## 1 Introduction

Despite commendable progress in expanding access to effective prevention and treatment interventions over the last twenty years, HIV/AIDS remains a significant public health concern particularly, in sub-Saharan Africa with approximately 25.6 million people living with HIV (PLWH) (1). Mental illness is a common comorbidity in PLWH due to the shared and intersecting vulnerabilities with HIV/AIDS (2). The presence of mental illness is associated with impaired judgment (3) which impedes timely and regular access to HIV prevention interventions (2). In addition, mental illness and stressful life events are associated with poor adherence to antiretroviral treatment and accelerated HIV disease progression characterized by CD4 cell count decline, increased viral load, and an elevated risk for clinical decline and mortality (4). Globally, there is a higher burden of mental health problems among PLWH compared to the general population (2,4,5). This pattern is attributed to the psychological distress associated with being diagnosed with a serious illness, a high burden of opportunistic infections, medication side effects, as well as the social stigma and discrimination associated with HIV (4,6).

Suicidality: a condition that refers to a wide spectrum of potentially harmful thoughts, behaviors, and experiences that often precede a fatal suicide attempt, is one of the major mental health problems associated with HIV (7). Suicidality occurs along a continuum of severity characterized by transient suicidal thoughts that progress to persistent ruminations about ending one’s life, development of concrete suicide plans, engaging in acts of intentional self-harm, and attempted suicide (8,9). These may occur independently or together with other psychiatric comorbidities such as major depressive disorder (MDD) (10).

Suicidal ideation is a predictor of future suicidal attempts and completed suicide (11,12), and PLWH are one hundred times more likely to commit suicide compared to the general population (13). A recent systematic review and meta-analysis study on suicidal ideation, attempts, and its associated factors among PLWH in Africa, reported a pooled prevalence of 21.7% (16.8-26.63%) for suicidal ideation and 11.06% (6.21-15.92%) for suicidal attempt (3). The substantially high lifetime prevalence of suicidal ideation and attempts is among PLWH (13) underlines the crucial need for tools that support the timely and accurate identification of people at risk of suicide. Suicide risk assessment is essential to suicide prevention and achieving health equity for PLWH(14) and should be a priority in PLWH, especially for those with more advanced disease (13).

Suicidality is a complex, multifactorial, and polygenic mental health problem that results from a variable combination of genetic, environmental, and behavioral risk and protective factors (15–18), each having small but meaningful contributions (19). These include situational factors such as a psychiatric diagnosis, hopelessness, impulsivity, perceived burdensomeness, stressful life events, social support, self-esteem, stable employment, problem-solving, and sense of belonging as well as static or non-modifiable factors such as gender, ethnicity, and psychiatric history (15). The sociodemographic, psychological, and clinical correlates of suicidality among PLWH in Uganda have been extensively studied and include low socioeconomic status (18,20,21), unemployment (20), lack of social support (22), stigma (20,21), poor problem-solving skills (22), state anger, trait anger, hopelessness, low self-esteem (23), low resilience (20), an increasing number of negative life events (11), past psychiatric history (11), anxiety symptoms (23), and MDD (11,21,23). Evidence from a genetic variation study among PLWH in Uganda implicated the S_A_ allele at the *5-HTTLPR/rs25531* locus in the serotonin transporter gene to be associated with increased suicidal risk (24). However, results from genome-wide association studies (GWAS) indicate that the genetic architecture of suicidal behavior is complex and highly polygenic (25–27) and recent GWAS findings have confirmed significant shared genetic heritability of suicidal behavior across ancestries (27–30).

The vast number and complexity of the risk factors associated with suicidal thoughts and behaviors limit the magnitude of statistical association with any single risk factor (31) rendering the prediction of suicidal behavior a complex classification problem that requires algorithms that can simultaneously consider tens or hundreds of risk factors to model complex relationships (32). Traditional statistical modeling approaches analyze a limited number of predictor variables at a time and cannot consider and account for complex and contingent interactions among risk factors (31). However, evidence from previous studies indicates that even well-known risk factors of suicidality have modest predictive strength individually (33). Therefore, suicidality predictive models developed using this approach perform only slightly better than random guessing (34).

Machine learning offers new tools to overcome challenges for which traditional statistical methods are not well-suited (35) because ML algorithms can process high-dimensional datasets, recognize complex patterns across multiple interacting risk factors (31), and determine the optimal model (36). As a result, ML has emerged as a promising tool for predicting future suicidal behavior (19,31,36,37) to support the timely identification of at-risk patients whose suicidality might otherwise have gone undetected (37).

Several ML approaches have been applied in the prediction of suicidal behavior. For instance, Nordin et, al. (37) identified eight different ML techniques commonly applied in the study of suicidal behavior, i.e. Bayesian-based approaches such as Naïve Bayes (NB), instance-based approaches, artificial neural network (ANN), regularization, decision tree (DT), support vector machine (SVM), regression, and ensemble learning techniques such as random forest (RF). In another systematic review of ML and the prediction of suicide in psychiatric populations, Pigoni et, al. (36) reported that random forests (RF), support vector machines (SVM), and convolutional neural networks often outperformed other algorithms. However, none of the studies included in the analyses were conducted in Uganda or among PLWH. Moreover, most previous studies did not incorporate genetic predictors and lacked external validation samples to evaluate prediction models.

We evaluated the potential of ML in predicting suicidality using a longitudinal data set of PLWH from Uganda, computed suicidality PRS for a subset of these participants, and examined whether incorporating genomic data could improve the predictive performance of suicidality prediction models trained using sociodemographic, clinical, and psychosocial data only.

## 2 Materials and Methods

### 2.1 Study design

This retrospective case-control study used sociodemographic, clinical, and genetics data collected between May 2012 and December 2013 by the European & Developing Countries Clinical Trials Partnership (EDCTP) funded Senior Fellowship Study. The EDCTP study was a prospective cohort study that investigated risk factors for psychiatric disorders among adults with HIV/AIDS in Uganda.

### 2.2 Study participants

The primary EDCTP Fellowship study recruited anti-retroviral therapy-naïve PLWH who were already enrolled in chronic HIV care at two specialized HIV clinics in Entebbe (semi-urban) and Masaka (rural) areas of Uganda. Participants were recruited in the study if they were at least 18 years of age and fluent in English or Luganda (the language in which the study instruments were translated) and were assessed for suicidality, MDD, and other psychiatric disorders at baseline and 12 months using the Mini International Neuropsychiatric Interview (M.I.N.I.) (38) based on the 4^th^ edition of the Diagnostic and Statistical Manual of Mental Disorders ( DSM-IV). Study participants who were too ill or unable to understand the study instruments and those who had missed their most recent scheduled clinic visit were excluded. All participants consented to future genetics research and provided blood specimens for DNA extraction. However, genetic data was obtained on only a subset of 282 participants.

### 2.3 Data description

The data set consists of baseline sociodemographic, psychosocial, and clinical assessment data on 1,126 participants, and clinical and psychosocial assessment data on 1,070 study participants at follow-up. In addition, we had individual-level genome-wide data on a subset of 282 participants generated using the H3Africa SNP array version 2, which accounts for the larger genetic diversity and smaller haplotype blocks in African genomes (39). The sociodemographic variables include age, sex, highest educational attainment, religious affiliations, marital status, food security, and employment status. In addition, participants provided information on whether they owned or rented their house, the type of construction material used for constructing the house occupied, whether they had access to electricity, and if they owned durable household assets such as a car, bicycle, radio, telephone, refrigerator, flask, and cupboard. The psychosocial variables included data on social support and the number of negative life events obtained using structured and standardized locally translated psychosocial assessment instruments which have previously been used among PLWH in Uganda (11,40). The clinical assessment variables include the duration of HIV infection, CD4 count, height, weight, HIV-related symptoms, HIV-associated neurocognitive impairment, and social impairment. The psychiatric assessment variables included data on previous psychiatric diagnoses, family history of psychiatric disorders, MDD, and suicidality based on the diagnostic output of MINI (38).

### 2.4 Data preprocessing

We cleaned the data and performed various mathematical and statistical transformations to convert the raw data into formats suitable for use in ML. For the religious affiliation, we reduced the number of independent categories by merging all participants with Christian-leaning beliefs into one category labeled as ‘Christians,’ and those with Islamic beliefs as ‘Muslims.’ Similarly, we merged the detailed subcategories of employment status into two categories, i.e., ‘employed’ for those with any form of employment or ‘unemployed, for participants without any form of employment. We computed each participant’s wealth index as a proxy measure of the socioeconomic status of a household socioeconomic status (SES) index by combining responses to questions on housing characteristics and ownership of eight durable household items commonly found in a typical Ugandan household using multiple correspondence analysis (41–44).

We obtained the psychosocial impairment index by summing responses to three questions on how HIV-related illness has disrupted normal activity in the past month with a higher score indicating higher impairment. Using the European Parasuicide Interview Schedule (EPIS) as modified by Kinyanda and colleagues (2005) for the Ugandan context (40), we derived the social support index, negative life events score, and stress score index. The social support index was obtained by summing responses to the items of the social support module of the modified EPIS. To generate data for computing negative life events score, participants were asked to report whether they had experienced any listed adverse life events in the past 6 months, and for each reported negative life event, participants rated how stressful it was on a 3-point Likert scale (45). We obtained the negative life events score by counting positive responses and the stress score index by summing the ratings of how stressful the reported adverse life events were.

To minimize the adverse impact of extreme values on the performance of our ML outcomes, we performed outlier detection to exclude participants whose age was 1.5 times higher or lower than the interquartile range. We categorized the remaining participants into five age groups based on an age interval of 10 years.

To obtain the binary target variable we coded all study participants who met the suicidality diagnostic criteria on MINI as cases and the rest were regarded as controls. We further categorized cases as prevalent if first diagnosed at baseline, incident if first diagnosed at follow-up, or persistent if diagnosed at both the baseline and follow-up. This coding yielded 207 prevalent cases with 919 controls, and 54 incident cases, with 1,016 controls. The data is characterized by the existence of a minority and majority class and is technically referred to as imbalanced data (46).

### 2.5 The problem of class imbalance in machine learning persistent

Machine learning algorithms work optimally with data in which the distribution of the number of instances is almost equal across the classes (46,47). Training models on imbalanced data can lead to biased models that fail to capture important patterns in the data (46) leading to prediction bias and poor performance of the model in the minority class (48). In suicidality prediction models, this poor performance could translate to missed opportunities to avert death by suicide. Therefore, we applied cost-sensitive learning (CSL) to improve classifier performance. Cost-sensitive learning alleviates the imbalanced data problem by assigning a higher cost for misclassifying the minority (positive) class (49). This strategy is not only computationally efficient, but it also preserves the data distribution (46).

### 2.6 Feature selection

The baseline data set consists of 300 variables including data on sociodemographic characteristics, HIV-related symptoms, HIV clinical status, and responses to a series of psychosocial and psychiatric assessment scales for 1,126 study participants. Similarly, the follow-up dataset includes 263 variables covering HIV clinical status, and psychosocial and psychiatric assessment interviews for 1,071 study participants. However, some variables are considerably redundant while others are irrelevant to the classification problem.

We selected suicidality predictors previously documented among PLWH in Uganda. To minimize collinearity between the selected predictor variables, we excluded one from each pair of predictors if their Pearson’s correlation coefficient (continuous) or Cramer’s V (categorical) was greater than 0.5. The final set of predictor variables for inclusion in the model was then selected using the least absolute shrinkage and selection operator (LASSO) algorithm. LASSO is a penalized regression algorithm that selects training features by gradually shrinking the coefficients of the less important features towards a mean of zero.

This process yielded a total of fourteen composite predictor variables including study site, sex, highest educational attainment, marital status, employment status, psychiatric history, MDD status, social support, stress, socioeconomic status, age, duration of HIV diagnosis, social impairment, and HIV-related dementia.

#### 2.6.1 Model development

We split the resulting data into training and test sets allocating 80% of the data for training and 20% for testing. We opted for an 80:20 split ratio because of the high number of predictor variables relative to the total number of available data points (50). To remedy the issue of missing data, we imputed missing values by replacing missing continuous data with the median value for the column and missing categorical data with the most frequent value for the column. We then transformed the updated data into features that better represent the underlying problem to the ML algorithms by scaling and normalization of continuous variables or one-hot encoding for categorical variables. The ML models were developed in Python version 3.11.9 (51) using supervised ML algorithms available in the Python library Scikit-learn version 1.3.1 (52)

#### 2.6.2 Machine learning approaches

We trained and evaluated eight classification algorithms frequently encountered in suicidal behavior prediction (37) to explore and select ML approaches that best capture the patterns in our data set. These include stand-alone algorithms such as logistic regression (LR), SVM, NB, k-nearest neighbors (KNN), and DT classifiers, as well as ensemble algorithms such as RF, adaptive boosting (AdaBoost), and gradient boosting (GB) classifiers.

Logistic regression is ideal for modeling linear relationships and is the most widely used algorithm for binary classification, due to its simplicity, and efficiency in handling large datasets. However, it struggles with modeling complex non-linear data. Support vector machines are more suited for the classification of non-linearly separable data. They perform classification in a single decision step by leveraging kernel functions to map non-linearly separable data into higher dimensional space to find a decision boundary (hyperplane) that best separates the different classes.

Naïve Bayes is a fast classification algorithm based on Baye’s theorem of conditional probability. It is robust in handling categorical features but is undermined by the assumption of strong conditional independence among predictor variables. On the other hand, the KNN algorithm assigns equal importance to all features, and it relies on the similarity of the training examples to the test data to predict the cluster to which a new object belongs by majority vote between the k-nearest neighbors.

Decision trees perform classification by recursively partitioning data into nodes and leaves creating interpretable tree-like structures. Random forests are an extension of decision trees that use majority voting to combine decisions from multiple decision tree models obtained from different subsets of the same dataset to produce a final classification decision. They are highly effective in processing high-dimensional, correlated data, and are considered the state-of-the-art algorithm in suicidality prediction. Another extension of the DT model is AdaBoost which refines predictions by iteratively focusing on misclassified examples, to produce a weighted ensemble model. The GB model is a robust, interpretable classification algorithm based on a gradient descent-based approach. It is particularly suited for handling noisy or incomplete data making it valuable in psychiatric research.

#### 2.6.3 Model performance

The performance of prediction models was assessed using a suite of classification metrics, including accuracy, F1-score, area under the receiver operating characteristic curve (AUC), sensitivity, specificity, positive predictive value (PPV), negative predictive value (NPV), and Mathew’s correlation coefficient (MCC). Each of these metrics is derived from the confusion matrix and conveys specific information about the quality of a classification (53). Accuracy is an intuitive metric depicting the overall proportion of correct predictions. However, it can be misleading in class-imbalanced datasets because it disregards the model’s performance on the minority class and provides an over-optimistic estimate of the classifier’s ability (54). Sensitivity and specificity are key measures of diagnostic accuracy that reflect the model’s ability to correctly classify positive and negative instances, respectively. Sensitivity and specificity are independent of disease prevalence but can vary depending on the spectrum of the disease in the studied group (55). Positive predictive value and NPV focus on the correctness of positive and negative predictions, offering a more nuanced understanding of prediction quality. The F1-score is the harmonic mean of precision(positive predictive value) and recall(sensitivity) and is a popular metric for imbalanced data sets. However, it can be misleading because it does not consider negative instances correctly classified by the ML classifier and is not invariant to class-swapping. The Pearson-Mathews correlation coefficient (MCC) relies on information from all four quadrants of the confusion matrix, and it is considered one of the most useful performance metrics for a binary classification in class-imbalanced data (56). The ROC curve AUC summarizes the model’s ability to discriminate between positive and negative cases across all decision thresholds. It is regarded as the most robust metric for comparing model performance because it is invariant to class imbalance (57).

### 2.7 Model optimization

We performed an exhaustive grid search across each model’s hyperparameter space using 10-fold stratified cross-validation with AUC as the scoring metric. The best parameters for tuning the models were selected using the mean cross-validation AUC across the ten folds.

### 2.8 Model explanation

We used the Shapley additive explanation (SHAP) approach to identify and visualize the important features contributing to the learning and prediction of the models. SHAP is a model-agnostic framework that offers a straightforward and consistent approach to interpreting model predictions by leveraging game theory techniques to assign a value (SHAP value) to each feature for a particular prediction (58). The mean absolute SHAP values were presented as a bar plot illustrating the relative importance of these input features for the models’ predictions.

### 2.9 Suicidality polygenic risk scores

To calculate suicidality PRS and determine if incorporating them in the selected baseline models improves their performance, we used individual-level genotype data for the subset of 282 study participants. Polygenic risk scores were computed using the clumping and thresholding (C+T) approach in PRSice-2 (59). Clumping and thresholding is a two-step process involving the selection of the most significant variants by pruning redundant correlated effects caused by linkage disequilibrium (LD) between variants followed by the removal of variants with a *p*-value larger than a chosen level of significance (*p* > *p*_T_) (60,61). The C+T method is preferred because it is intuitive, requires few assumptions, and is relatively easy to implement (61).

#### 2.9.1 Target data preparation

The genotype data were converted to Plink binary format using PLINK 1.9 (62) and then subjected to rigorous pre-and post-imputation quality control (QC). The pre-imputation QC steps were performed using the human heredity and Health Africa (H3A) GWAS pipeline (63) and included the removal of duplicate SNPs, individuals with discordant sex information, non-autosomal SNPS, SNPs with minor allele frequency (MAF) < 0.05, poorly genotyped SNPs with genotyping rate < 0.9, poorly genotyped individuals with SNP missingness >0.02, and SNPs that violated the Hardy-Weinberg equilibrium (HWE) p-value threshold of 1e^-6^. In addition, we performed relatedness, and heterozygosity checks to exclude closely related pairs of individuals based on identity by descent (IBD) ≥ 0.11 and samples with high heterozygosity ≥ 0.34 or low heterozygosity ≤ 0.15. Overall, 1,780,439 SNPs and 262 samples passed the pre-imputation QC filters and were used for SNP imputation. The untyped SNPs were imputed against the Africa Genome Resources (AGR) reference panel on the Sanger Imputation Service (64). In the post-imputation QC, each of the 22 chromosome files was processed separately to exclude SNPs with MAF < 0.01, imputation info score (INFO) < 0.8, and HWE *p*-value <1 e^-6^. The quality-controlled data was merged into a single file containing 12,420,057 SNPs for 261 EDCTP study participants.

#### 2.9.2 Base data

The base data was derived from European ancestry GWAS summary statistics consisting of 8,905,379 SNPs on the full-scale ordinal suicidality scale in the UK Biobank cohort (25). After standard GWAS QC to exclude SNPS with INFO < 0.8 and MAF < 0.01, we retained 8,904674 SNPs.

### 2.10 Ethical considerations

This research obtained ethical approval from the Makerere University, College of Health Sciences, School of Biomedical Sciences Institutional Review Board under reference number SBS-2023-473.

## 3 Results

### 3.1 Characteristics of the study participants

Of the 1,126 study participants with baseline data, 207 (18.4%) had a positive suicidality diagnosis based on the B items of the MINI. These were coded as cases, while the remaining 921 participants were coded as controls. The majority (94.7%) of the participants who met the suicidality diagnostic criteria scored less than 9 on the B items of the MINI (Fig1), implying that most participants had low severity of suicidality symptoms. The distribution of participants by severity of suicidality symptoms is provided in Figure 1.

**Figure 1:**
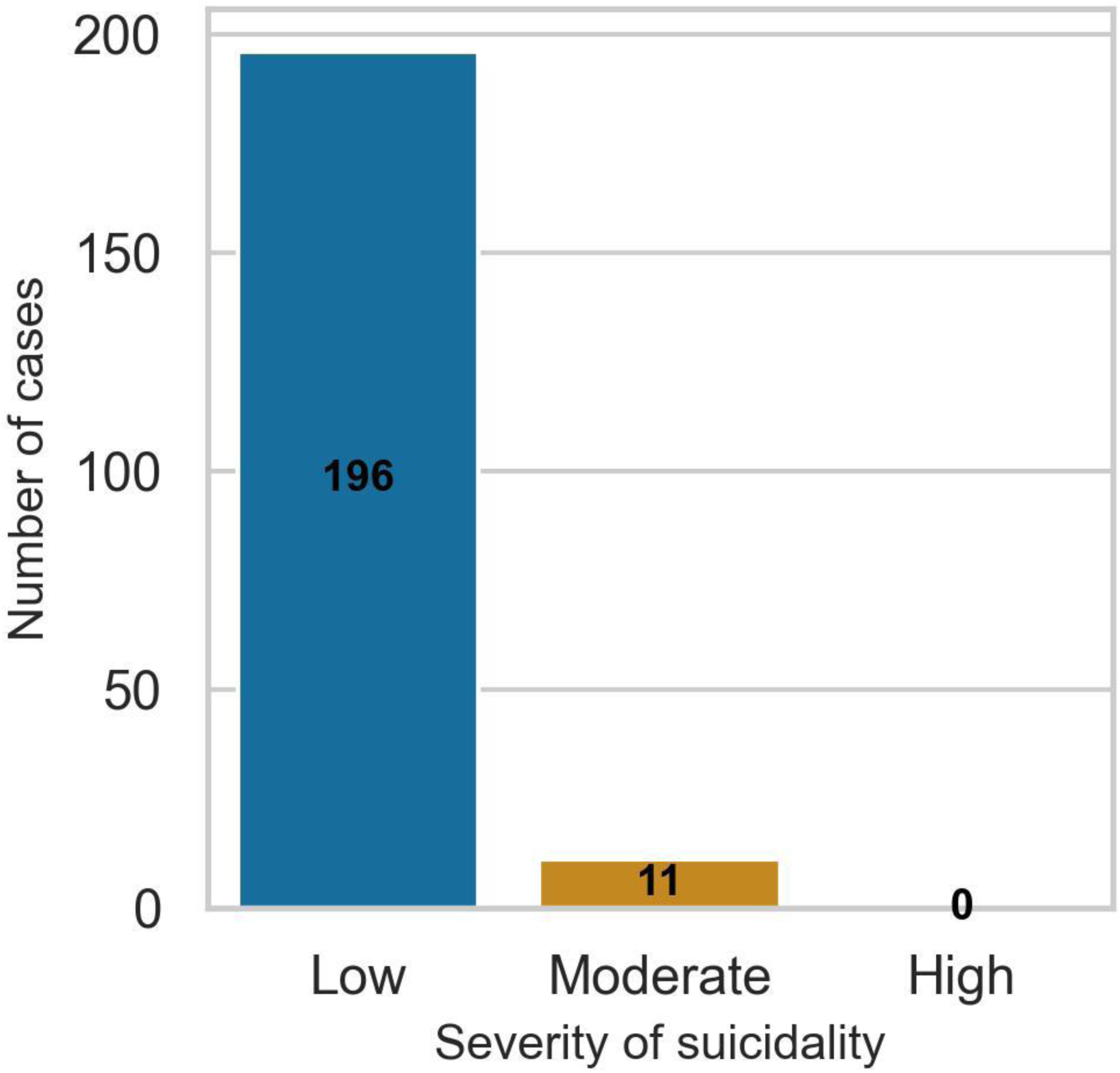
Bar chart for the distribution of study participants by the severity of their suicidality symptoms at baseline. Suicidality categories are based on the B items of the MINI with cut-off scores of <9 (low), 9 to 15 (moderate), and >16 (high).

Most of the study participants (77.2%) were female, aged between 18 and 82 years with a mean age of 35 (SD 9.3) years, had attained primary-level education (61.8%), and were married (51.5%). A total of 17 participants whose age was 1.5 times higher or lower than the interquartile range were excluded from further analysis. The final models were developed using data from 1,109 participants including 205 cases, and 904 controls. At follow-up, 56 (5.0%) of participants were lost to follow-up leaving 1,070 participants of whom 86 (8.0%) had a positive suicidality diagnosis. The demographic characteristics of all the study participants are presented in Table 1.

**Table 1:** Baseline demographic characteristics of EDCTP study participants disaggregated by enrolment site.

| Study variable |  | Studsit1(Entebbe) | Studsit2(Masaka) | Total<br>N(%) |
| --- | --- | --- | --- | --- |
| Sex | Male | 128 | 129 | 257 (22.8) |
|  | Female | 437 | 432 | 869 (77.2) |
| Educational<br>attainment | None | 54 | 70 | 124 (11.0) |
|  | Primary | 306 | 389 | 695 (61.7) |
|  | Secondary | 187 | 91 | 278 (24.7) |
|  | Tertiary | 17 | 9 | 26 (2.3) |
|  | Missing | 1 | 2 | 3(0.3) |
| Marital<br>status | Married | 304 | 276 | 580 (51.5) |
|  | Widowed | 50 | 114 | 164 (14.6) |
|  | Separated/Divorced | 135 | 136 | 271 (24.1) |
|  | Single | 75 | 34 | 109 (9.7) |
|  | Missing | 1 | 1 | 2 (0.2) |

### 3.2 Model selection and optimization

We trained and evaluated eight ML models using default parameters. All the models performed better than random guessing, achieving overall AUCs between 0.59 and 0.78 with low sensitivity and high specificity. Table 2 shows the comparative performance of the baseline suicidality prediction models across selected binary classification metrics. The GB, AdaBoost, LR, and RF demonstrated slightly better discriminative ability, achieving an overall AUC of 0.78, 0.77, 0.76, and 0.73, respectively. On the other hand, NB and DT achieved greater sensitivity compared to the other models.

**Table 2:** Performance of ML algorithms for predicting suicidality with default parameters.

|  | Accuracy | AUC | MCC | Sensitivity | Specificity | F1-score |
| --- | --- | --- | --- | --- | --- | --- |
| **RF** | 0.84 | 0.73 | 0.37 | 0.29 | 0.97 | 0.41 |
| **GB** | 0.83 | 0.78 | 0.33 | 0.27 | 0.96 | 0.37 |
| **AB** | 0.81 | 0.77 | 0.27 | 0.29 | 0.93 | 0.36 |
| **NB** | 0.75 | 0.72 | 0.24 | 0.44 | 0.82 | 0.40 |
| **LR** | 0.81 | 0.76 | 0.22 | 0.22 | 0.94 | 0.30 |
| **KNN** | 0.82 | 0.64 | 0.21 | 0.15 | 0.97 | 0.24 |
| **DT** | 0.76 | 0.59 | 0.18 | 0.32 | 0.86 | 0.33 |
| **SVM** | 0.82 | 0.69 | 0.16 | 0.07 | 0.99 | 0.13 |

RF: random forest, GB: gradient boosting, AB: AdaBoost, NB: naïve Bayes, LR: logistic regression,
KNN: k-nearest neighbors, DT: decision tree, SVM: support vector machines.

Figure 2 shows the comparative discriminative performance of the classification models during training and testing phase.

**Figure 2:**
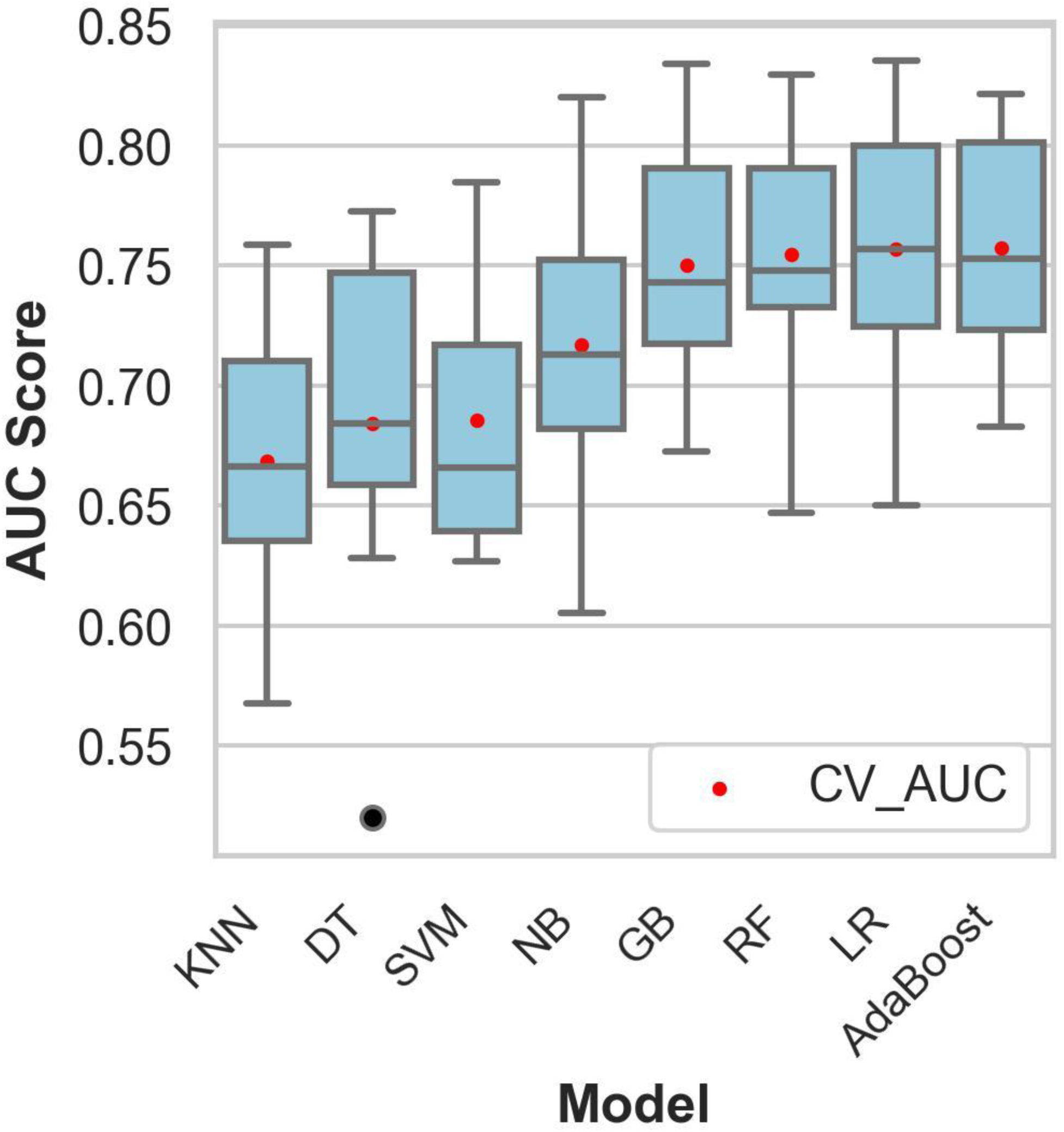
Comparative performance of the models on the training dataset. The mean cross-validation AUC was obtained using stratified 10-fold cross-validation. CV_AUC: mean cross-validation area under the curve, KNN: k-nearest neighbors, DT: decision tree classifier, SVM: support vector machines, NB: naïve Bayes, GB: gradient boosting, RF: random forest, LR: logistic regression.

After hyperparameter tuning, the AdaBoost and GB models performed similarly, achieving an overall AUC of 0.79 on the test dataset. However, inspection of the combined ROC curves (Figure 3) revealed multiple points where the two curves intersect, implying that either model could have greater sensitivity than the other for some specificity thresholds.

**Figure 3:**
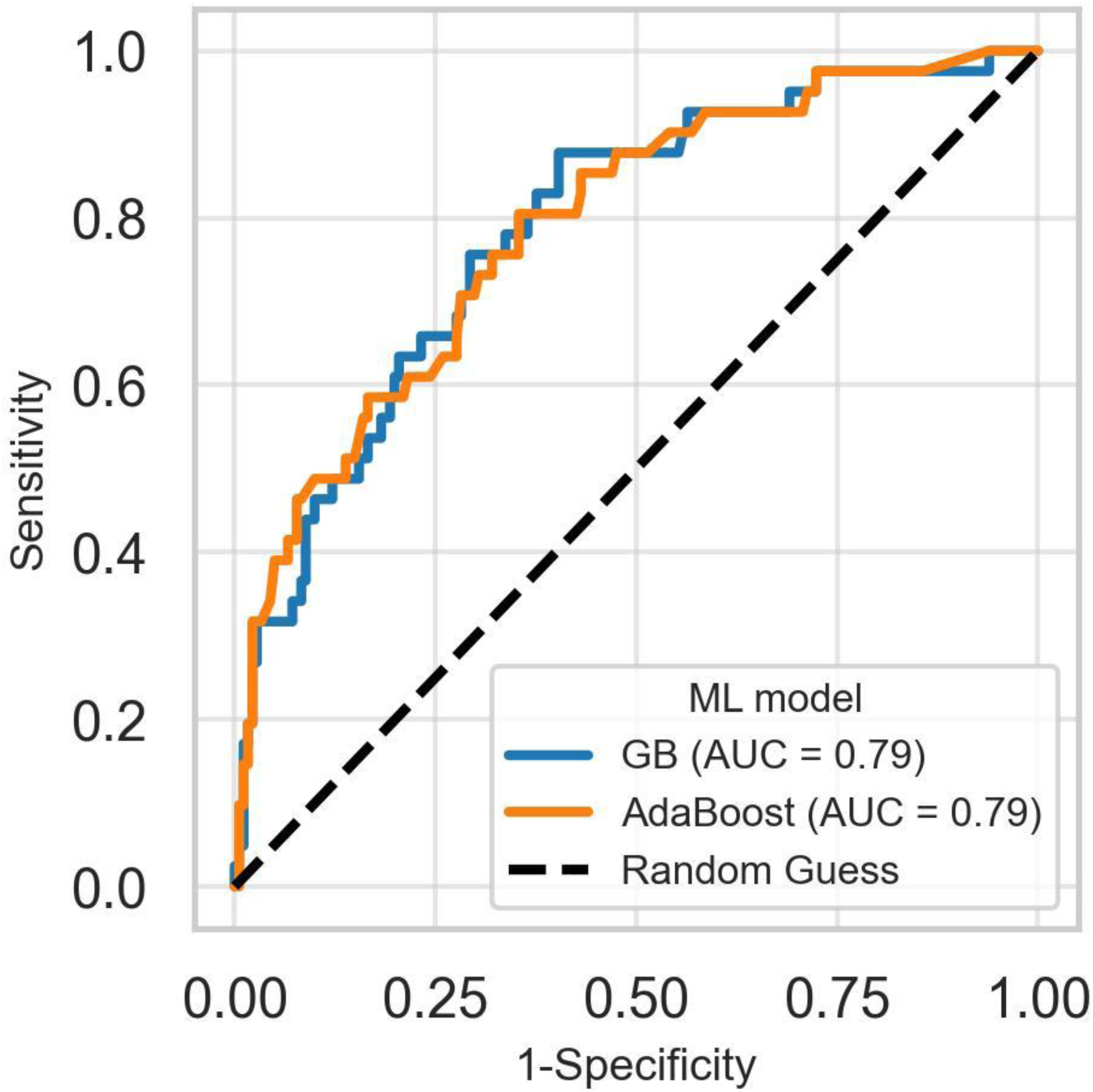
Receiver operating characteristics curves of the best-performing models for predicting suicidality. The ROC curves for AdaBoost and GB models intersect at several points making it difficult to choose the best-performing model based on overall AUC. GB: Gradient boosting classifier

Since overall AUC is computed by integrating the model’s performance across all possible thresholds, selecting a superior model when two ROC curves intersect is challenging. To overcome this impasse, we computed the models’ partial AUC focusing on the ROC curve’s early retrieval (ER) area, which shows the model’s ability to correctly identify positive cases when the false positive rate (FPR) is low. The partial AUC corresponding to an FPR of 0.1 ( ROC-AUC_0.1), revealed that AdaBoost was the best model for predicting suicidality. Figure 4 shows McClish’s corrected partial AUC at an FPR of 0.1 and 0.2 corresponding to misclassification of 10% and 20% of negative cases as positive.

**Figure 4:**
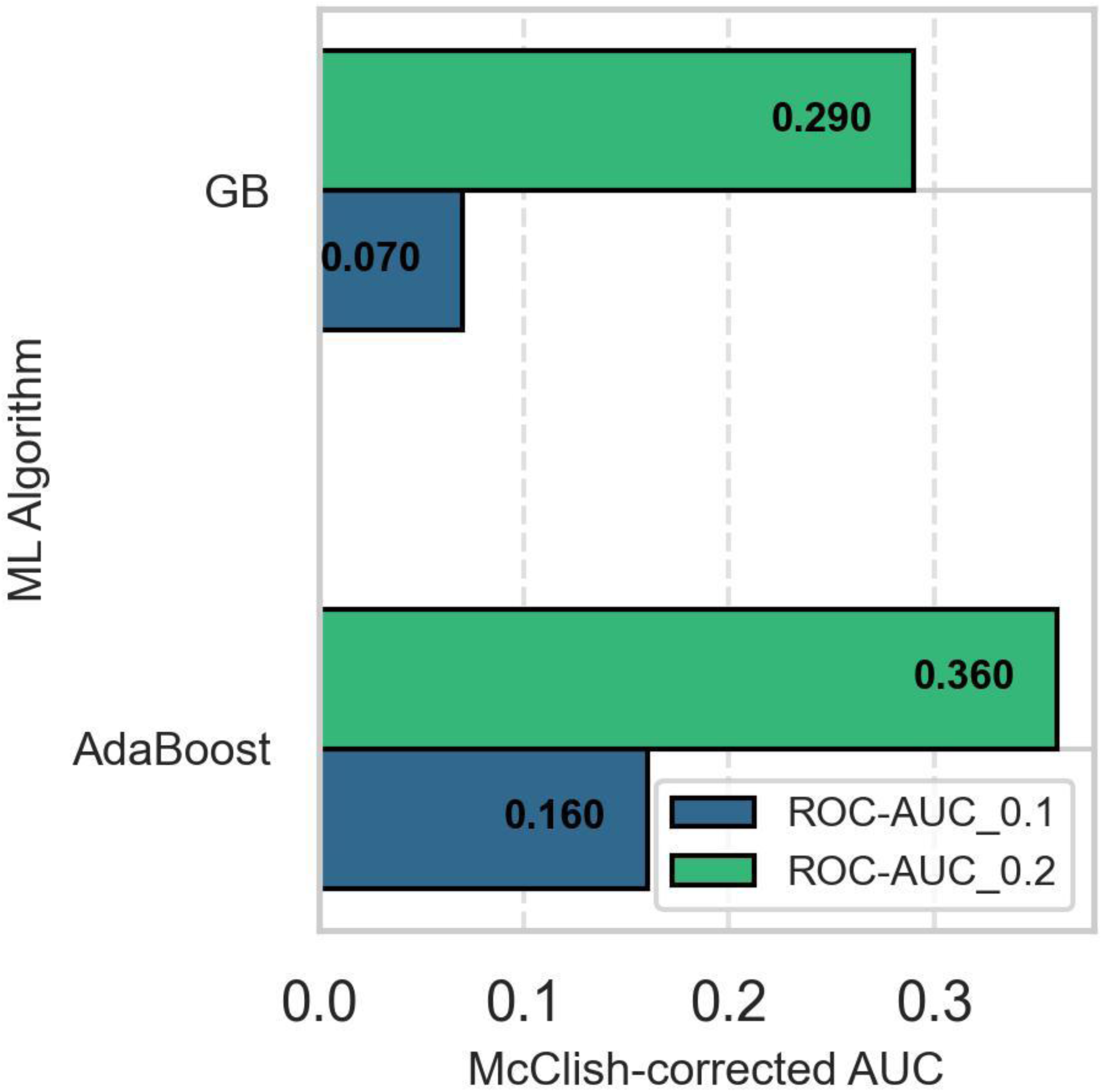
Comparison of McClish-adjusted early retrieval AUC of the AdaBoost and GB models. AdaBoost model is superior to GB at predicting the positive class when the false positive rate is low. ROC-AUC_0.1: area under the receiver operating characteristics curve when FPR is 10%, ROC-AUC_0.2: area under the receiver operating characteristics curve when FPR is 20%.

### 3.3 Model performance after correcting the class imbalance

Using the class imbalance ratio of 4.44 as the penalty for the misclassification of the minority class, we developed a cost-sensitive AdaBoost model that achieved a sensitivity of 0.63, a specificity of 0.74, a PPV of 0.36, and an NPV of 0.89. A summary of the model performance before and after correcting class imbalance is provided in Table 3.

**Table 3:**
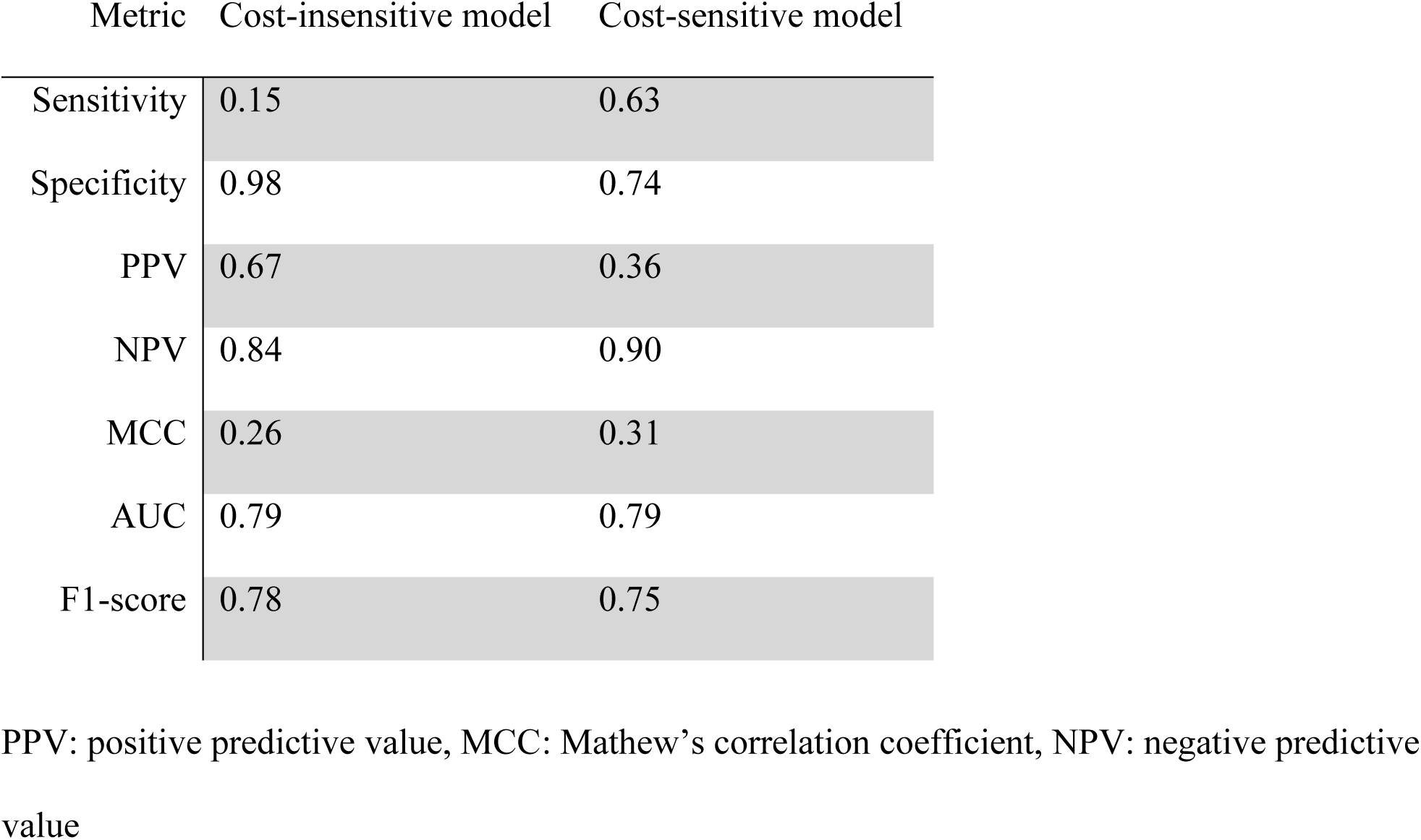
Comparative performance of cost-insensitive and cost-sensitive AdaBoost model for predicting suicidality.

The most important features contributing to predictions in the model were a negative MDD diagnosis, low-stress category, positive MDD diagnosis, HIV dementia, high-stress category, low psychosocial impairment, moderate social support, being aged 28–37 years, being married, and having low social support. The features contributing to predictions in the AdaBoost model are shown in Figure 5.

**Figure 5:**
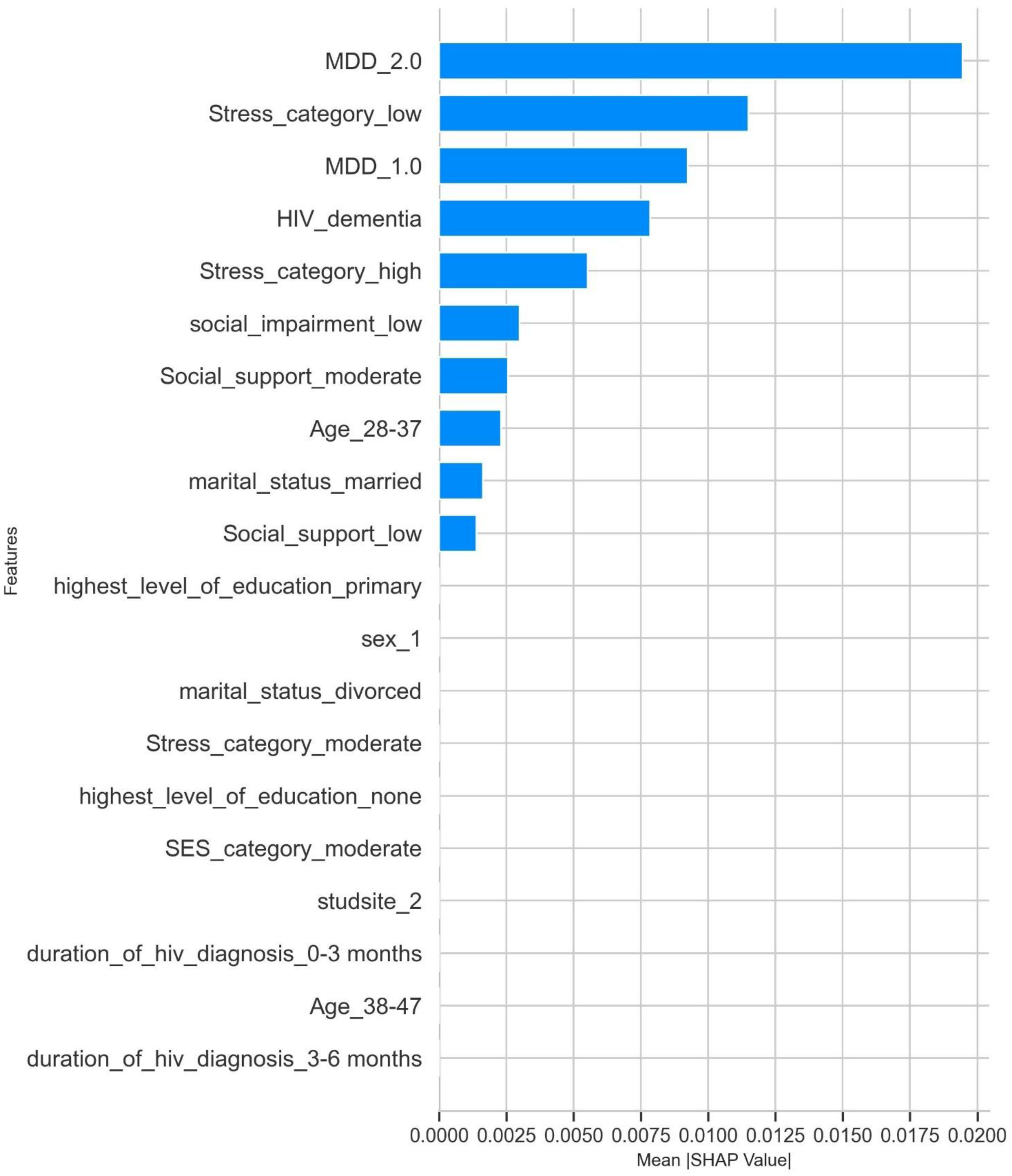
Features contributing to suicidality predictions in the cost-sensitive AdaBoost model. MDD_1.0: positive MDD diagnosis, MDD_2.0: negative MDD diagnosis, studsite2: Masaka (rural), sex_1: male, SES: socioeconomic status.

### 3.4 Predicting future suicidality risk

To evaluate the generalizability of our prediction model, we validated its performance using psychosocial and clinical data collected at the 12-month follow-up. The cost-sensitive AdaBoost model achieved an overall AUC of 0.75 (95% CI: 0.69 - 0.81), a sensitivity of 0.41, a specificity of 0.93, a PPV of 0.33, an NPV of 0.95, and MCC of 0.31 in predicting 12-month suicidality risk. In addition, the model predicted incident suicidality with an AUC of 0.69 (95% CI: 0.62 - 0.76), a sensitivity of 0.33, a specificity of 0.91, a PPV of 0.17, and an NPV of 0.96.

### 3.5 Suicidality polygenic risk scores

There were 360 shared SNPs between the discovery and the target data. The best-fit PRS was obtained from effect size estimates of the shared SNPs at a p-value threshold of 0.0002 and was statistically significant (*p*-value 0.007). In addition, a standard two-sample t-test revealed a statistically significant difference (*p*-value 0.008) between the population means of the PRS of cases and controls. To estimate the variance explained by suicidality PRS, we fitted a mixed effects logistic regression model in R (65) and calculated the Nagelkerke pseudo-R^2^, with and without the best-fit PRS as the full and null model, respectively. After adjusting for the covariates of sex, age, MDD status, and the first ten principal components, suicidality PRS accounted for 3.3% of the phenotypic variance between cases and controls.

### 3.6 Impact of incorporating suicidality polygenic risk scores in the prediction model

We trained and evaluated the performance of a cost-sensitive AdaBoost model using the baseline data of the 282 participants for whom individual-level genetic data were available. The model achieved an overall AUC of 0.79 (95% CI: 0.66 – 0.91), with a slightly higher sensitivity (0.75), PPV (0.41), and MCC (0.39) compared to the model trained using sociodemographic, psychosocial, and clinical predictors only. Figure 6 shows the most prominent features contributing to suicidality predictions in the model trained using a combination of sociodemographic, clinical, psychosocial, and genetic risk factors.

**Figure 6:**
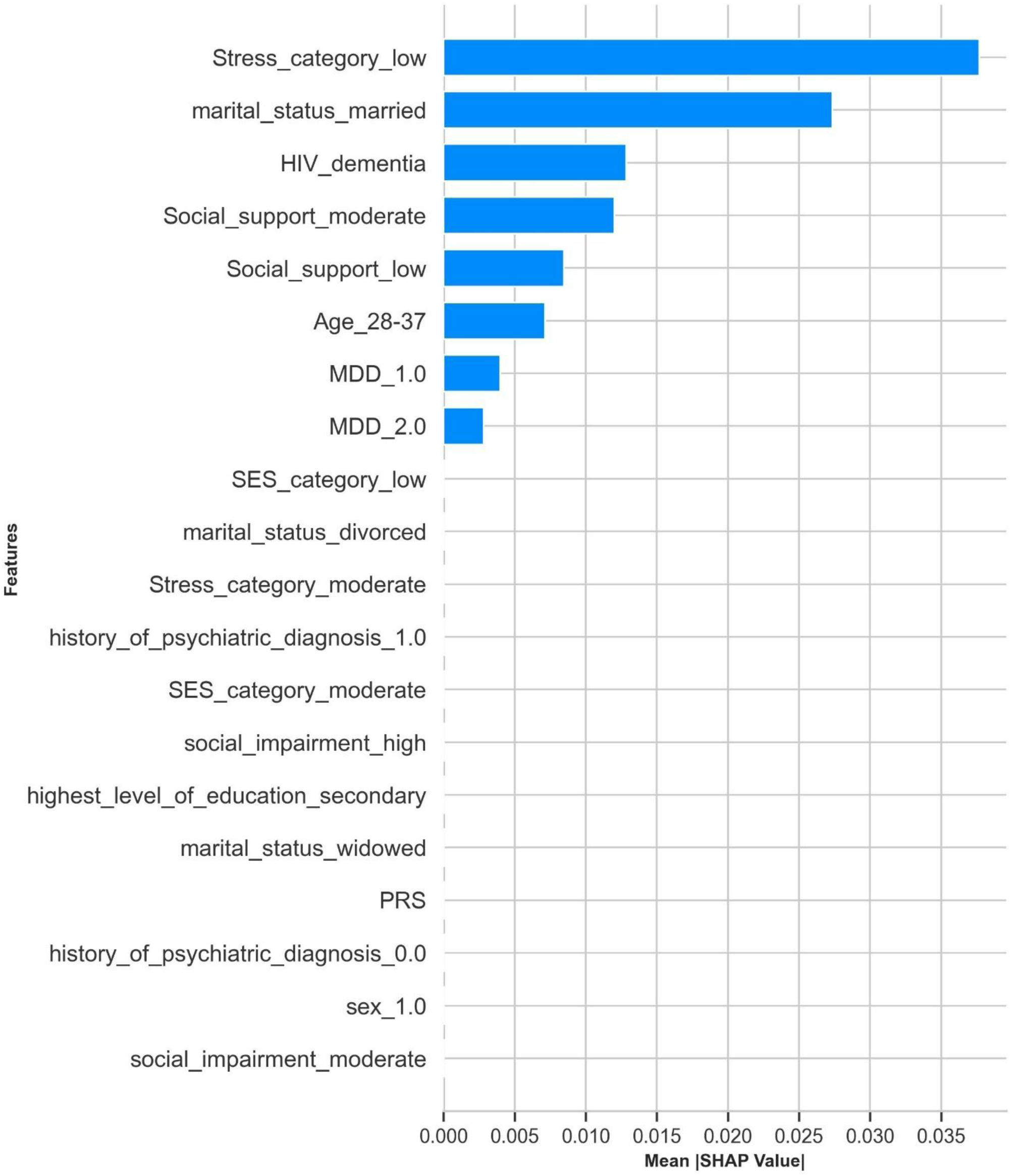
Feature importance of the variables contributing to suicidality prediction after incorporating PRS in the cost-sensitive AdaBoost model. MDD_1.0: positive MDD diagnosis, MDD_2.0: negative MDD diagnosis, PRS: polygenic risk score, SES: socioeconomic status, sex_1.0: male, sex_2.0: female, history of psychiatric diagnosis_1.0: positive previous psychiatric diagnosis, history of psychiatric diagnosis_0.0: no previous psychiatric diagnosis

## 4 Discussion

This study explored the potential of various ML algorithms in predicting suicidality using longitudinal sociodemographic, clinical, and psychosocial data of PLWH in Uganda. In addition, we computed suicidality PRS and assessed their contribution to the predictive performance of the models. To the best of our knowledge, this is the first study to apply machine learning to predicting suicidality in PLWH in Uganda. In addition, it is the first study to integrate genetic and environmental risk factors into an ML framework for suicidality prediction in PLWH within Africa.

Overall, models developed using ensemble learning algorithms outperformed the stand-alone models across all the selected binary classification metrics with an overall AUC ranging between 0.72 and 0.79 across the top five prediction models. The AUC is a summary metric of the ROC curve with values ranging between 0 and 1, and according to the proposed ranges for interpreting the relationship between AUC and the diagnostic accuracy of the model (55), our models are considered ‘very good’ at distinguishing between diseased and non-diseased individuals. Nonetheless, even our best-performing model would be of limited clinical utility (66).

We employed cost-sensitive learning to correct class imbalance and improve model performance while preserving the underlying distribution of the data. Cost-sensitive learning resulted in a 320% increment in model sensitivity and a 24% and 46% decline in specificity and PPV, respectively. According to Wang et al (2021), sensitivity and PPV are functions of the model cut-off threshold, such that an increase in sensitivity is accompanied by a rapid increment in the FPR that in turn results in a reduced PPV(67).

Regarding the importance of features measured using SHAP values, a negative MDD status, low-stress levels, positive MDD diagnosis, HIV dementia, high-stress levels, low psychosocial impairment, moderate social support, age between 28 and 38 years, and being married were among the top ten features contributing to suicidality predictions. A formal MDD diagnosis and high stress constituted the most significant positive predictor of suicidality risk.

Major depressive disorder is a major risk factor for suicidality among PLWH (11,20,68,69) and the general population (15,70). In the same vein, stressful life events are recognized as triggers of suicidal behavior, and it is believed that exposure to chronic stress can progressively erode an individual’s resilience in dealing with stressful situations and lead to a higher probability of engaging in suicidal behavior (71). These findings are consistent with the stress vulnerability model of suicidality which asserts that suicidal behavior involves vulnerability or diathesis as a distal risk factor that predisposes individuals to suicidal behavior when stress is encountered (72).

To our knowledge, there have been no previous studies that used ML for suicidality prediction among PLWH, however, our results are consistent with the findings from studies in the general population. Su *et, al* (73) developed a model for predicting self-harm and suicide attempts using data on 2,809 Australian adolescents and an AUC of 0.74 and 0.72 for self-harm and suicide attempts, respectively. In addition, they reported that depression was the most important factor for predicting self-harm and suicide attempts (73). Similarly, Bazrafshan *et, al.* (74) reported an AUC of 0.8 for an RF model developed using longitudinal data on a sample containing 3,833 cases obtained from hospitals across the Illam Province in Iran. They also reported that age group, educational level, marital status, and employment, among others, were significantly associated with suicide (74). On the other hand, Macalli *et, al.* (75) reported an AUC of 0.80 in predicting suicidal thoughts and behaviors among 5,066 college students in France.

We obtained a statistically significant best-fit PRS (p-value 0.007) despite the low number of shared SNPs between the base and target data. While the small number of shared SNPs can be explained by the differences in the LD patterns of the discovery (European ancestry GWAS) and the target (African ancestry), the statistically significant PRS score implies that the selected SNPs are strongly associated with suicidality and is a testament to the shared genetic heritability of suicidality across ancestral populations (26–29).

On the other hand, the variance explained by suicidality PRS was low accounting for 3.3% of the phenotypic differences between suicidality cases and controls. However, this is not surprising because the predictive power of PRS decays with increasing genetic distance of the target cohort from the discovery cohort. Incorporating PRS as an additional feature in the model resulted in a 19%, and 14%, improvement in model sensitivity and PPV and a 4% reduction in specificity.

## 5 Strengths and limitations

The key strength of our study is that we utilized longitudinal data, consisting of both dynamic and static risk factors for suicidality. We relied on a cost-sensitive learning approach to mitigate the class imbalance in the data by assigning a higher misclassification cost for the positive class. In addition, we incorporated suicidality PRS data in our model to account for the contribution of genetic variation in suicidality risk. We also evaluated the generalizability of our models by predicting suicidality at a 12-month follow-up.

A key limitation of our study is that we did not perform model calibration yet poorly calibrated risk predictions could result in unnecessary and costly interventions or missed opportunities potentially causing patient harm (76). Another limitation of our study relates to the broad definition of suicidality phenotype used in developing the model. Considering that the severity of suicidality among the study participants was generally low, it might prove difficult to justify additional resources for preventive interventions.

## 6 Conclusion and recommendations

Machine learning models can effectively predict suicidality using the sociodemographic, psychosocial, and clinical data of PLWH in Uganda. This is significant because these data are easy to collect and are readily available in electronic format in most HIV care and treatment centers in Uganda. The ROC-AUC_0.1 depicts the model’s ability to predict the positive class enabling a straightforward comparison and selection of a suitable model in scenarios where minimizing false positives is critical. Incorporating suicidality PRS improves the overall predictive performance of the model developed using sociodemographic, psychosocial, and clinical predictors.

Larger studies that include more diverse participants are required to validate whether the inclusion of suicidality PRSs in clinical prediction models can enhance the stratification of patients at risk of suicide attempts. In addition, future studies could be designed to assess the utility and cost-effectiveness of deploying suicidality prediction models in a clinical setting

## Data Availability

All data produced are available online at https://github.com/abmutema/Predicting-suicidality-in-people-living-with-HIV-in-Uganda

## 10 Code availability

All the code used in this analysis is available at: https://github.com/abmutema/Predicting-suicidality-

### in-people-living-with-HIV-in-Uganda

## 12 Author Contributions

Conceptualization: AK, EK and ABM, Data curation: ABM

Formal analysis: ABM Methodology: ABM, AK Supervision: AK, DJ, EK Writing -original draft: ABM

Writing -review and editing: LL, AK, DJ, EK, SF Funding: EK

## 14 Acknowledgments

